# U-shaped effects of humidity and the impact of misspecified weather-transmission relationships in epidemic models

**DOI:** 10.64898/2026.10.05.26364750

**Authors:** Cana Kussmaul, Pietro Gemo, Laura Andrea Barrero Guevara, Matthieu Domenech de Cellès

## Abstract

Understanding the relationship between weather conditions and infectious disease transmission has important public health implications; nevertheless, the functional form of this relationship often remains poorly understood. Humidity has previously been suggested to have a U-shaped effect on the transmission of respiratory pathogens, with optimal transmission occurring at both low and high levels. Here, to assess the impact of this functional form and of its potential misspecification, we first simulate transmission rates based on the U-shaped relationship and then test whether an exponential function accurately estimates these rates across a range of climates. We further use a transmission model to simulate case count data for a range of hypothetical pathogens, comparing outbreak dynamics under the U-shaped forcing function to those under the best-fit exponential approximation. In our simulations, the model fit for the exponential function is climate-dependent, performing best in very dry and very humid environments. In locations where specific humidity values centre around the vertex of the U-shaped function, the exponential model underestimates the amplitude and frequency of incidence peaks. These findings demonstrate the potential consequences of assuming an incorrect functional form for weather-infectious disease relationships and highlight the need for more careful consideration of this functional form in modelling practice.

## Main

Accurate models of how weather affects the transmission of infectious diseases are central to understanding and predicting the dynamics of disease outbreaks and therefore have important public health implications (1,2). One potential source of inaccuracy for such models is the assumed functional form of the relationship between a given weather variable and disease transmission. For respiratory pathogens, various measures of humidity have been shown to have an effect on transmission (2–6). While this relationship has often been modelled with an exponential or otherwise monotonic forcing function (7), more recent evidence suggests that a U-shaped function, i.e., with high transmission during both low and high humidity, better describes the relationship for a range of respiratory pathogens (2,6,8).

An implication of the U-shaped functional form is that an annual periodicity in the forcing variable can yield twice-yearly peaks in the transmission rate, since, if the values of the forcing variable range across both arms of the U-shape, high transmission will occur during both its peak and its trough. This contrasts with monotonic functional forms, which would imply annual peaks in the transmission rate in this scenario. While disease incidence does not necessarily follow the same trajectory as the transmission rate, more frequent peaks in the transmission rate may also impact the frequency of peaks in incidence.

Here, we use specific humidity (SH) as an example to 1) investigate the impact of assuming an incorrect functional form for weather-transmission relationships and 2) examine the potential consequences of a U-shaped effect on the periodicity of transmission case counts. To this end, we obtained SH data from cities representing a wide range of climates. We used this data and a U-shaped function (2) to generate the ground-truth effect of SH on the transmission rate for each city. We then used a regression model to fit an exponential function of SH to approximate these transmission rates. This process functions as a computationally inexpensive proxy for fitting a full transmission model to simulated incidence data. Finally, using a transmission model, we simulated case count data for both the fitted exponential function and the ground-truth U-shaped function.

As the first step of our analysis, we compared the goodness-of-fit of the exponential model to the “true” transmission rate (based on a U-shaped function) across different climates. A clear pattern emerges when plotting model fit against the mean SH in each city (Figure 1A): in locations with a mean SH close to the centre of the underlying U-shape (fixed at 11 g/kg in our simulations), the fit of the regression model assuming an exponential relationship is drastically worse compared to locations with a more extreme mean SH. This is because cities where SH values centre around ∼11 g/kg (e.g., Tokyo and Houston) tend to span both arms of the U-shaped function, making it difficult for the necessarily monotonic exponential function to capture the true relationship. The resulting regression coefficients for the strength of forcing in these scenarios are very small and statistically insignificant (Table 1), demonstrating that assuming the wrong functional form here also leads to an underestimation of the humidity effect and reduced sensitivity to detect an effect at all. Locations that are consistently very dry (e.g., Reykjavik) or very humid (e.g., Taipei), on the other hand, only “make use” of one side of the U, making it much easier to approximate the effective function with an exponential (Figure 1B).

**Table 1.** Model fit (*R*^2^) and strength of forcing coefficient (*δ*_*sH*_) for the regression models with an exponential term fitted to the ground-truth transmission rate.

| Location | Mean SH<br>(g/kg) | R<sup>2</sup> | $\delta_{SH}$ | Standard error<br>for $\delta_{SH}$ | p-value for<br>$\delta_{SH}$ |
| --- | --- | --- | --- | --- | --- |
| Reykjavik | 4.8 | 0.99 | -0.071 | 0.001 | <0.0001 |
| Taipei | 15.4 | 0.98 | 0.232 | 0.001 | <0.0001 |
| Timbuktu | 7.2 | 0.93 | -0.341 | 0.003 | <0.0001 |
| Houston | 12.1 | 0.04 | 0.004 | 0.033 | 0.8929 |
| Tokyo | 10.1 | 0.01 | 0.001 | 0.072 | 0.9901 |

**Figure 1.**
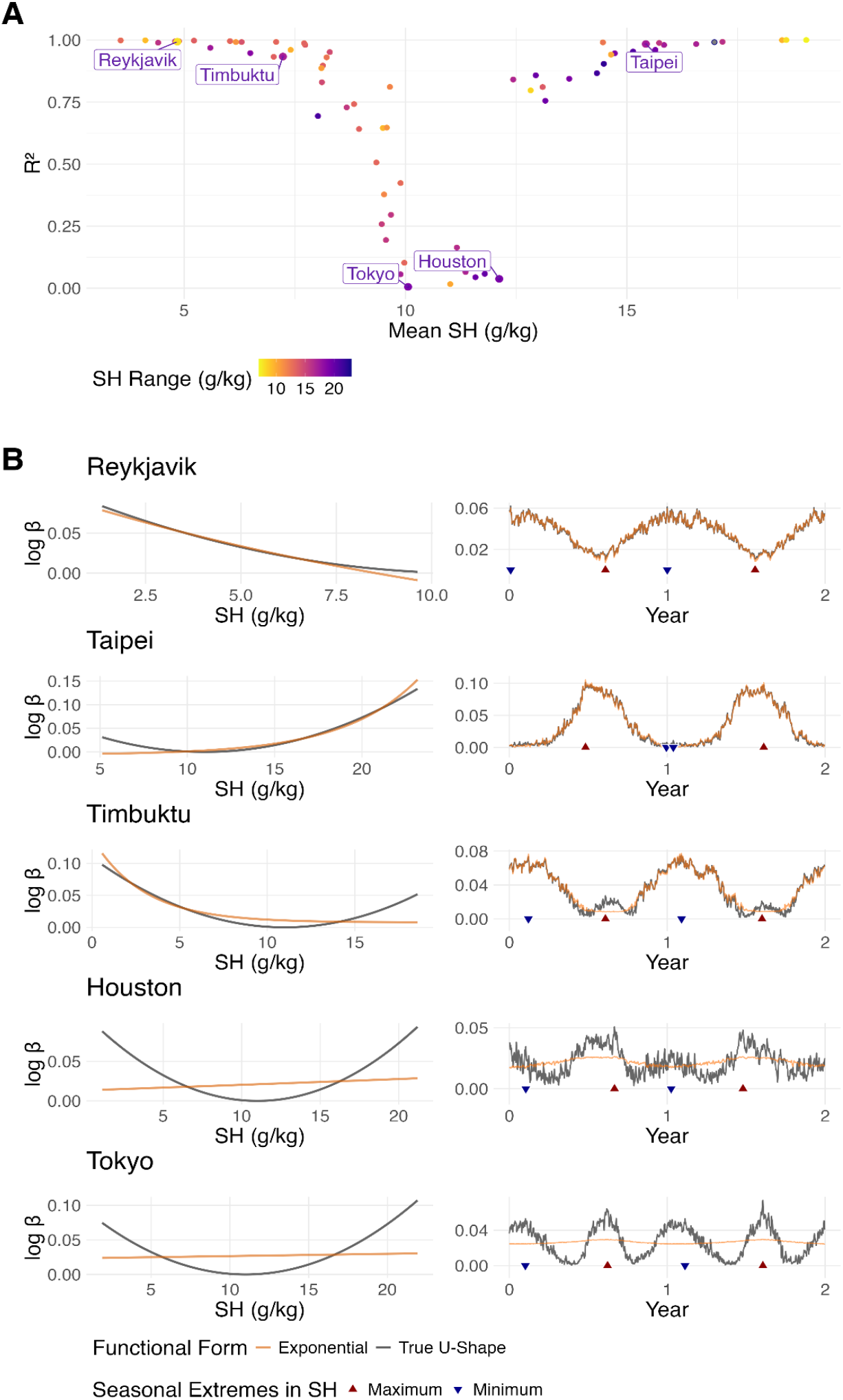
A: Regression model fit (*R*^2^) by mean SH and SH range for 70 cities. B: Ground-truth U-shaped functional form for the effect of SH compared to the fitted exponential function for the range of SH observed in each city (left) with the resulting 2-year seasonal pattern of the transmission rate (triangles indicate minimum and maximum SH-values in the locationspecific seasonal pattern of SH) (right).

An additional consequence of the U-shaped function is that the forced transmission rate can exhibit two peaks when the forcing variable peaks only once per season. For our five focal locations, we find that whether and to what extent this occurs again depends on the range of specific humidity values represented in a given location. The second peak becomes more pronounced the more the specific humidity values span the full U-shape. When it does occur, the exponential model predictably fails to capture this second peak in the transmission rate (see Tokyo, Houston, and Timbuktu; Figure 1B).

In the second step of our analysis, we assessed how these differences in the transmission rate translate to disease incidence. We extracted coefficients from the regression models fitted in step one and simulated case count data with both the original U-shaped forcing function and the fitted exponential approximation for the five focal cities. To cover a range of hypothetical respiratory pathogens, we also simulated across a grid of parameter values for the duration of immunity and the mean age at first infection. We found that, on average, cases peaked at a lower frequency when the mean age at first infection was high — a result expected based on epidemiological theory (9).

While it is important to note that the patterns observable in our simulations are not a necessary consequence of either functional form, any differences in case count patterns between the equivalent U-shape- and exponential-based simulations can be attributed to the functional form, since all other factors are kept constant. As a result of the exponential function successfully approximating the U-shaped forcing function in these cities, simulations for Taipei and Reykjavik exhibited no notable differences in case count patterns between the assumed functional forms. However, this is not the case for Houston and Tokyo, where SH values span the centre of the true forcing function, and the regression fit was poor as a result. In these cities, two important differences can be observed. Firstly, since the best exponential approximation of a perfectly symmetrical U-shape is a horizontal line, the effect of SH (as extracted from the exponential regression model) is very small in Tokyo and Houston. The resulting case count therefore oscillates at a far lower amplitude than that based on the U-shape, leading to a mismatch between each of the two equivalent time series (Supplementary Figure S1).

Secondly, the periodicity of the simulated case count differs between the U-shaped model and the exponential approximation for Houston and Tokyo. Simulations based on the exponential model resulted in very little variation in periodicity, with the vast majority of the wavelet power concentrated at the annual period for both cities (Figure 2). This is also reflected in the dominant period, which was annual across the entire parameter space. Equivalent simulations based on the U-shape, on the other hand, had dominant periodicities ranging from 0.5 (i.e., semi-annual peaks) to 1.4 years, with all simulations exhibiting both annual and semi-annual patterns. Notably, the dominant period in Tokyo was semi-annual for 28.9% of U-shape-based simulations, including for some with a high mean age at first infection — a property that leads to lower-frequency outbreaks in the simulations for all other locations. Overall, this suggests that while periodicities in case count depend on several factors, the functional form of the weather effect can also play an important role. The double peak in the transmission rate resulting from the U-shaped forcing function can lead to a range of incidence patterns, including but not limited to sub-annual periodicities.

**Figure 2.**
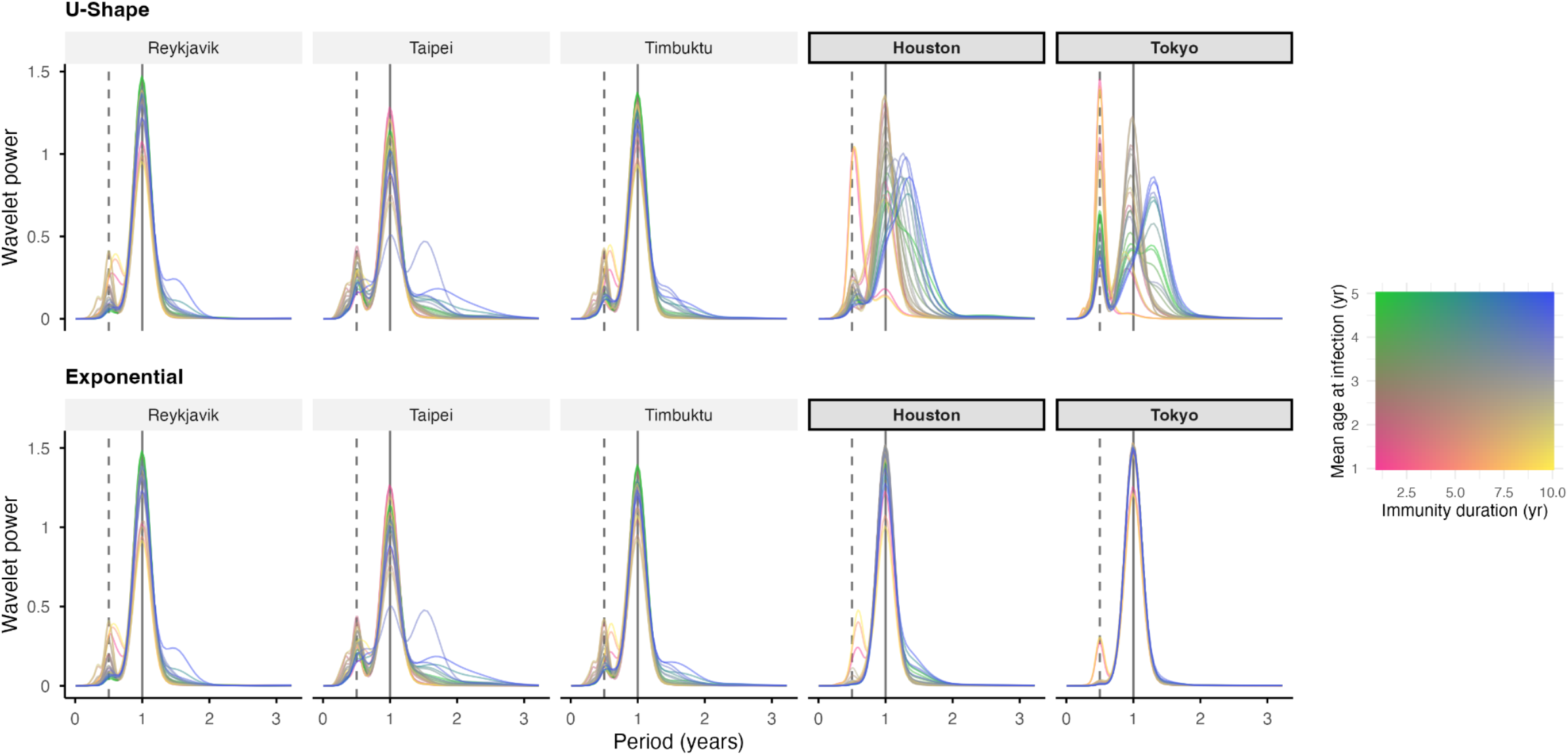
Wavelet power by city and period for simulated case count data. Each curve is a single simulation with the colour indicating the position in the parameter space. The solid vertical line marks an annual period, and the dashxed line a 0.5-year period.

In summary, we show that if the true underlying functional form for the effect of a weather variable, such as specific humidity, is U-shaped, the impact of assuming an exponential forcing function differs by climate: if a climate is such that humidity values are restricted to a single side of the U-shape, the choice of functional form has little impact on the model fit nor on the resulting dynamics in incidence. Conversely, when humidity values span a substantial range both above and below the vertex of the U, the fit of an exponential forcing function is poor, and the importance of the humidity effect is likely to be underestimated. Consequently, in such locations, the implied trajectory of disease incidence differs substantially across assumed functional forms, with the exponential model often underestimating both the amplitude and frequency of peaks in cases. Thus, we caution against using a monotonic function as a default in this context and instead encourage considering more complex functional forms to accurately capture the relationship between meteorological variables and disease transmission. Due to word count constraints, we discuss the study’s limitations in the supplementary materials.

## Methods

In this simulation study, we aimed to investigate the potential impact of U-shaped humidity effects on the periodicity of transmission and incidence, and more generally, the impact of assuming an incorrect functional form for the effect of weather variables. We did this in two stages: First, we generated a ground-truth transmission rate based on a U-shaped effect of SH and fitted a regression model with an exponential term to this transmission rate. At this stage, we compared the model fit across climatically diverse locations. Second, we extracted the fitted parameters for the exponential model and used an SE_2_I_2_RS model to simulate case count data, based on the fitted exponential and the true U-shape, across a range of locations and parameter values. We then used wavelet analysis to compare epidemic dynamics between the models. Details, including model equations, are available in the supplementary materials.

### Step 1: Defining and fitting functional forms

The functional form for the effect of humidity on transmission, serving as the ground-truth for this study, was roughly based on a function described by Baker et al. 2019 for the effect of specific humidity on RSV transmission (2). We assumed a quadratic function with the minimal transmission potential at an SH of 11 g/kg. For each location, we calculated the resulting “true” seasonal transmission rate using 15 years of specific humidity data (2010-2025). To assess the ability of the exponential to approximate the patterns caused by the U-shaped function, we then fitted a nonlinear least-squares model with an exponential term to the transmission rates for each location and used R^2^ as a goodness-of-fit metric.

### Step 2: Simulating and comparing case count data

We specified a fully deterministic SE_2_I_2_RS model (see supplementary materials). The force of infection in this model is modulated by the transmission rates as calculated in step 1. We simulated 36 years of case count data for each functional form, each of the five focal locations, and a range of synthetic pathogens. Hypothetical pathogens were simulated by sweeping over a grid of parameter values for the mean age at first infection and the average duration of immunity D. The mean age at first infection ranged from 1 to 5 years (in 0.5-year increments), and the mean duration of immunity ranged from 1 to 10 years (in 2-year increments). To keep locations and functional forms comparable, we ensured a constant time-averaged transmission rate at the same parameter values.

We then performed a wavelet analysis on the last 10 years of each simulated dataset and extracted the average power and period, as well as the dominant period, and compared the resulting periodicities across different functional forms and parameter values.

## Data Availability

All data and code will be made available upon publication in a peer-reviewed journal.

## Acknowledgements

This work was funded by the Max Planck Society. The authors gratefully acknowledge the members of the Domenech Lab at the Max Planck Institute for Infection Biology for a helpful group discussion of the manuscript.

