## Supplementary Materials for "U-shaped effects of humidity and the impact of misspecified weather-transmission relationships in epidemic models"

#### Supplementary Discussion

In our simulations, we assume the same U-shaped pattern identified for RSV applies to all hypothetical pathogens. While similar shapes have been identified, for e.g. influenza (1), this assumption likely does not hold across all respiratory pathogens. Whether or not a specific climate results in a successful approximation with a monotonic function depends on the shape and, if present, the vertex of the humidity effect. Hence, the climates identified as "problematic" here do not necessarily generalise, i.e., a monotonic function may fit fine in Tokyo and Houston for a different pathogen. As long as the true shape is non-monotonic, however, we expect the overall conclusions to hold: 1) whether or not a monotonic function can successfully approximate the true weather-transmission relationship depends on the climate in a given location, and 2) this can impact model fit and the implied periodicity of incidence. This also means our work may have implications for pathogens with entirely different non-monotonic relationships to different weather variables. For example, temperature and rainfall have been shown to have an inverted U-shaped effect on the transmission of vector-borne diseases (2–4), which may lead to similar phenomena.

Further, for computational efficiency, we fitted a regression model to the simulated transmission rate, rather than fitting a full transmission model to simulated incidence data. Assuming that all other parameters and the model structure are correctly specified, a transmission model would be expected to give essentially equivalent maximum likelihood estimates for the humidity effect to the regression we run, since the likelihoods maximised by these methods are linked (5). However, in a real model-fitting scenario, the poor fit we demonstrate here would likely be compensated for with other estimated parameters to some extent. This may result in a less obviously poor fit, but crucially, could lead to the misestimation of other parameters. With our approach, we isolate the impact of the functional form alone under otherwise ideal conditions.

Lastly, one aspect we aimed to address with this project was the potential of biennial peaks in transmission resulting from annual peaks in the forcing variable. We therefore chose to focus the more detailed analyses on 5 locations that each exhibit a regular annual pattern in SH. The

70 locations we ran the regression model for were not selected like this and instead represent a wider range of climate patterns. The general point about poor fit in climates that span both sides can therefore be expected to generalise to these climates, while the effect on the periodicity of peaks in transmission and incidence may differ.

Overall, this work is of a theoretical nature, intended to draw attention to the potential impact of an often-overlooked modelling choice. To this end, we have made a number of simplifying assumptions in our models and have not aimed to recreate any specific patterns observed in real incidence data. For example, we did not include school terms or weather variables other than specific humidity, our model is purely deterministic, and it does not include an observation model. Nevertheless, our results have real implications for modelling practice and, by extension, public health: by highlighting the need to carefully consider the assumed functional form of the effects of weather, we hope to contribute to improved estimates of these effects, more accurate models overall, and thereby improved guidance for public health interventions.

#### Data

We obtained daily specific humidity data (1989-2025), from the National Aeronautics and Space Administration (NASA) Langley Research Center's Prediction Of Worldwide Energy Resources (POWER) project. Data were extracted for 70 cities across the globe, with more detailed analyses focusing on 5 focal cities that vary in the range of SH values observed in their climates but each exhibit distinct annual peaks in the seasonality of SH: Houston, Reykjavik, Taipei, Timbuktu, and Tokyo.

#### Functional Forms

The ground-truth functional form for the effect of humidity on transmission was based on a function described by Baker et al. 2019 for the effect of specific humidity on RSV transmission (6):

$$\log(\beta_t) = (9 \times 10^{-4})\,(SH_t - 11)^2$$

The nonlinear least-squares model we fitted to the true $\log(\beta_t)$ can be described as follows:

$$\log(\beta_t) = \alpha + (e^{\delta_{SH}(SH_t - \nu)} - 1)$$

All parameters, i.e., the intercept $\alpha$, the forcing strength $\delta_{SH}$, and the shift parameter $\upsilon$, were estimated to make the exponential as flexible as possible.

#### Transmission Model

To simulate incidence data, we used an $SE_2I_2RS$ model described by the following difference equations initialised at their unforced endemic equilibrium.

$$S_{t+1} = S_t + \mu N - (\lambda_t + \mu)S_t + \omega R_t$$

$$E_{1,t+1} = E_{1,t} + \lambda_t S_t - (2\sigma + \mu)E_{1,t}$$

$$E_{2,t+1} = E_{2,t} + 2\sigma(E_{1,t} - E_{2,t}) - \mu E_{2,t}$$

$$I_{1,t+1} = I_{1,t} + 2\sigma E_{2,t} - (2\gamma + \mu)I_{1,t}$$

$$I_{2,t+1} = I_{2,t} + 2\gamma(I_{1,t} - I_{2,t}) - \mu I_{2,t}$$

$$R_{t+1} = R_t + 2\gamma I_{2,t} - (\mu + \omega)R_t$$

Where $\mu$ is the birth and death rate, $\omega$ the rate of waning immunity, $1/\sigma$ the average latent period, and $1/\gamma$ the infectious period. $\lambda$ is the force of infection, given by

$$\lambda_t = \beta_{seas}\frac{I_1 + I_2}{N}$$

The transmission rate, $\beta_{seas}$ , is the product of $\beta_t$ , the forcing function for specific humidity described above, and the baseline transmission $\beta_0$:

$$\beta_0 = \frac{R_0 (2\gamma + \mu)^2 (2\sigma + \mu)^2}{4(4\gamma + \mu)\sigma^2}$$

Since $\beta_0$ is constant across time and all variation in the transmission rate is due to $\beta_t$ , $\beta_t$ is referred to as the transmission rate throughout the manuscript for simplicity.

#### Rescaling $R_0$

To keep locations and functional forms comparable at the same parameter values, we rescaled $R_0$ by the mean forcing function ( $\bar{f}$ ) and used the rescaled version $R_0^{eff}$ to generate the data.

$$R_0^{eff} = \frac{R_0}{\bar{f}}$$

#### Numerical Implementation

All analyses were done in R 4.4.2. The transmission model was implemented in the package pomp (6.3). Non-linear regression models were fitted using the package minipack.lm (1.2-4). Wavelet analysis was performed using the WaveletComp (1.2) package. WaveletComp standardises input time series before they undergo wavelet transformation, enabling direct comparison across different time series since the resulting measure of wavelet power is relative to unit-variance white noise (7).

### Supplementary Figures

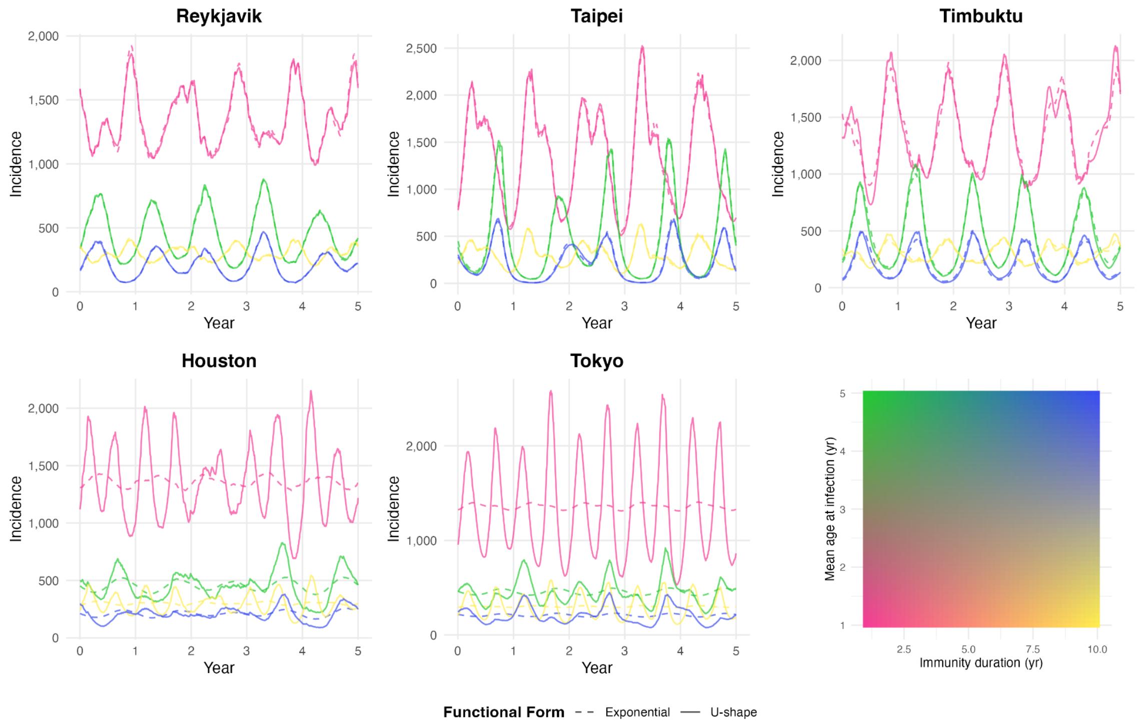


Figure S1. Incidence for the last 5 years of simulations by location and functional form. Each line is one simulation with the colour indicating the position on the parameter space. Four extremes of the parameter space are visualised here for better legibility.

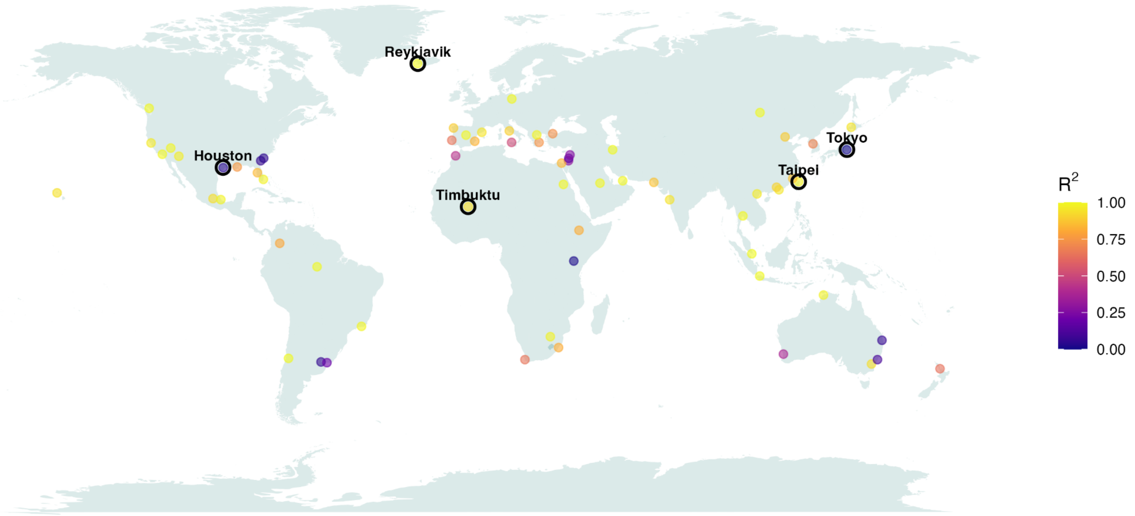


Figure S2. Map of study locations with the colour indicating regression model fit. Focal locations included in more detailed analyses are highlighted.
